# Study Protocol of A Phase II Study to Evaluate Safety and Efficacy of Neo-adjuvant Pembrolizumab and Radiotherapy in Localized Rectal Cancer

**DOI:** 10.1101/2021.10.31.21265647

**Authors:** Claudia Corrò, Nicolas C. Buchs, Matthieu Tihy, André Durham-Faivre, Philippe Bichard, Jean-Louis Frossard, Giacomo Puppa, Thomas McKee, Arnaud Roth, Thomas Zilli, Christelle Trembleau, Mariagrazia Di Marco, Valérie Dutoit, Pierre-Yves Dietrich, Frédéric Ris, Thibaud Kössler

## Abstract

**Background:** Reshaping the tumor microenvironment by novel immunotherapies represents a key strategy to improve the treatment of cancers. Nevertheless, responsiveness to these treatments is often correlated with the extent of the T cell infiltration at the tumor site. Remarkably, microsatellite stable rectal cancer is characterized by poor infiltration and, therefore, do not respond to immune checkpoint blockade. To date, the only available curative option for these patients relies on extensive surgery. With the aim to broaden the application of promising immunotherapies, it is necessary to develop alternative approaches to promote T cell infiltration into the tumor microenvironment of these tumors. In this regard, recent evidence shows that radiotherapy may have profound immunostimulatory effects, hinting at the possibility of combining it with immunotherapy. The combination of long-course chemoradiotherapy and immunotherapy was recently shown to be safe and yielded promising results in rectal cancer, however short-course radiotherapy and immunotherapy have never been tested in these tumors.

**Methods:** Our clinical trial investigates the clinical and biological impact of combining pembrolizumab with short-course radiotherapy in the neo-adjuvant treatment of localized microsatellite stable rectal cancer. This phase II non-randomized study will recruit 25 patients who will receive short-course preoperative radiotherapy (5Gy x 5 days) and four injections of pembrolizumab starting on the same day and on weeks 4, 7 and 10. Radical surgery will be performed after three weeks from the last pembrolizumab injection. Our clinical trial also includes an extensive research program involving the transcriptomic and proteomic analysis of blood and tumor samples throughout the course of the treatment.

**Discussion:** Our study is the first clinical trial to provide with safety and efficacy information of this novel treatment approach in rectal cancer, leading to a major breakthrough in the treatment of this cancer. Additionally, the translational research program will offer better insight into immunological changes within the tumor and blood during treatment. Taken together, our work will help optimizing future treatment combinations and, possibly, better selecting patients.

**Trial registration:** This study was registered with www.clinicaltrial.gov: NCT04109755. Registration date: June, 2020.

## Background

Rectal cancer (RC) is one of the most frequently occurring cancers and leading cause of cancer death worldwide [1-3]. Apart from surgery, which has a massive impact on the quality of life of RC patients, management of RC largely relies on conventional methods such as chemotherapy (CT) and radiotherapy (RT). For tumors localized close to the anal margin or node positive tumors, which represent 45 to 60% of the newly diagnosed patients, European and American oncology guidelines recommend either five weeks of neoadjuvant chemo-RT (CRT) followed by an 8- to 12-week treatment free interval before radical surgery, or a short-course preoperative RT (SCPRT) with surgery taking place either 1-2 weeks after RT or after 8 to 10 weeks [4, 5]. These two modalities of treatment are currently judged equivalent by the experts [6, 7].

The importance of intact immune surveillance function in controlling outgrowth of neoplastic transformations has been known for decades [8]. In recent years, the field of cancer immunotherapy has seen considerable progress due to the discovery of new therapies targeting immune checkpoint molecules such as cytotoxic T lymphocyte-associated protein 4 (CTLA-4), programmed cell death 1 (PD1) and programmed cell death ligand 1 (PD-L1) [9]. Clinical trials investigating these immune checkpoint inhibitors (ICIs) in advanced tumor settings have shown positive results in several cancers and these treatments have been integrated into the standard of care [10, 11]. Although the treatment with ICIs can provide deep and durable responses in some cancers, the clinical benefits are usually limited to a subset of patients. In gastrointestinal cancers, very impressive results have been obtained with an anti-PD1 antibody (pembrolizumab) in heavily pre-treated patients with advanced disease carrying mismatch repair deficient genes (dMMR) [12]. Tumors with dMMR are typically associated with microsatellite instability (MSI). Consequently, MSI-High (MSI-H) tumors are associated with a higher response rate to immunotherapies compared to microsatellite stable (MSS) tumors. This difference is thought to be due to a higher number of neo-antigens in MSI-H tumors compare to MSS tumors [13]. Recently, Overman and colleagues have published the result of an anti-PD1 immunotherapy in second line treatment in MSI-H metastatic colorectal cancer (CRC) [14]. These early results showed 50% progression free survival (PFS) at two years and 60% overall survival (OS) at two years, outperforming the results obtained with second line chemotherapies [14]. Unfortunately, MSI-H tumors represent only 1 to 3% of RCs. Therefore, the majority of RC are MSS and do not respond to immunotherapy.

Accumulating evidence shows a correlation between tumor-infiltrating lymphocytes (TILs) in cancer tissue and favorable prognosis in various malignancies [15]. In particular, the presence of CD8^+^ T cells and the ratio of CD8^+^ T cells/FoxP3^+^ regulatory T cells (Tregs) correlates with improved prognosis and long-term survival in solid malignancies [16]. As T cell infiltration is typically dictated by the presence of tumor-specific antigens (neoantigens), it is not surprising that MSI-H tumors display a greater number of TILs as compared to MSS cancers. In line with these observations, high “immunoscores” were found predictive of response to ICIs [17, 18].

In order to broaden the application of ICI to MSS RC, novel therapeutic approaches aiming at reconditioning the tumor microenvironment (TME) by either promoting TIL activation and infiltration are required. To this end, RT might be used, as recent evidence suggests that ionizing radiation can induce important immunomodulatory effects in the TME allowing the trafficking of T cells into the tumor [19]. These studies speak in the favor of combining RT and ICIs. Lately, a bulk of work in pre-clinical tumor models investigating the effect of radio-immunotherapy in many solid cancers, including CRC, highlighted the potential benefit of this approach [20-22]. Following these results, a few clinical trials are evaluating the combination of RT and ICIs in RC (NCT02948348, NCT04124601, NCT04262687, NCT04558684), but always in combination with CT and in advanced clinical settings. Encouraging safety and toxicity profiles from these studies indicate that radio-immunotherapy combinations could represent a valid opportunity for RC patients. For instance, in the VOLTAGE trial (NCT02948348) where patients received CRT followed by the anti PD1 antibody nivolumab, only mild toxicity was reported [23]. Moreover, 30% of patients with locally advanced MSS RC reached pCR. Remarkably, our clinical trial represents the first study where the impact of combining the anti-PD1 antibody pembrolizumab with short-course RT in the neo-adjuvant treatment of localized RC will be evaluated (NCT04109755). Alongside this clinical trial, a translational research project will provide a deep understanding of the dynamic changes in the immune and tumor cell states that are associated with responses to treatment and patient outcome. Ultimately, these results will have profound implications for the treatment of RC patients and the design of future clinical trial protocols for other non-inflamed tumors.

## Method and Design

### Objectives

The **primary objective** of this study is the assessment of the *Tumor Regression Grade (TRG)* after radio-immunotherapy. **Secondary objectives** include *Tolerability and Safety, OS, DFS, Locoregional Relapse-Free Survival (LRRFS), Distant Metastasis-Free Survival (DMFS), Quality of Life, Post-Operative Complication* and *Quality of Surgery*. Lastly, this clinical trial comprises an **exploratory objective** that overlap with a translational research program aiming at providing new insights into the tumor and immune cell states that are associated with response to treatment and identify novel prognostic and predictive biomarkers for clinical decision making.

### Study outcomes

The clinical trial will assess the following parameters:

1. *TRG* using the Mandard regression grade score. This parameter will be evaluated at the completion of the radio-immunotherapy treatment;
2. *Tolerability and Safety* using the common terminology criteria for adverse events (CTCAE) Version 4.0 and Dindo Clavien classification of surgical complication. These parameters will be evaluated at the completion of the radio-immunotherapy treatment;
3. *OS*, defined as the time from study entry until death due to any cause. Subjects who have not died at the time of last known follow-up will be censored;
4. *DFS*, defined as the time from study entry until recurrence, second primary cancer, or death without evidence of recurrence or second primary cancer;
5. *LRRFS*, defined as the time from surgery until local or regional recurrence. LRRFS is evaluated in patients who had an R0 resection only;
6. *DMFS*, defined as the time from surgery to metastatic recurrence;
7. *Quality of Life* using EORTC QLQ-C30 questionnaire and EORTC QLQ-CR29;
8. *Post-Operative Complication* according to the Dindo-Clavien score;
9. *Quality of Surgery* according to Nagtegaal’s recommendations;
10. *Tumor Immunome* and *Peripheral Immune Responses* in relation to the clinical outcomes.
  a. Comparison of the tumor immune microenviroment before (treatment-naïve) and after neo-adjuvant radio-immunotherapy using RNA sequencing and flow cytometry techniques.
  b. Quantitative and qualitative assessment of the immune responses in the periphery by analysis of peripheral blood (peripheral blood mononuclear cells (PBMCs), circulating tumor DNA (ctDNA), serum and plasma).
  c. Correlation of these findings with clinicopathological parameters and clinical outcomes.

### Study design

This trial is a phase II, single arm study, including 25 patients with localized RC. Following diagnosis and workout, patients will receive SCPRT (5Gy x 5 days) and four injections of pembrolizumab, on weeks 1, 4, 7 and 10. This phase of treatment will last 12 weeks, then surgery will be performed. Immune response will be monitored in the blood, specifically the peripheral blood mononuclear cell (PBMC) and within the tumor before, during radio-immunotherapy and after surgery. An outline of the study design is depicted in **Figure 1**.

**Figure 1.**
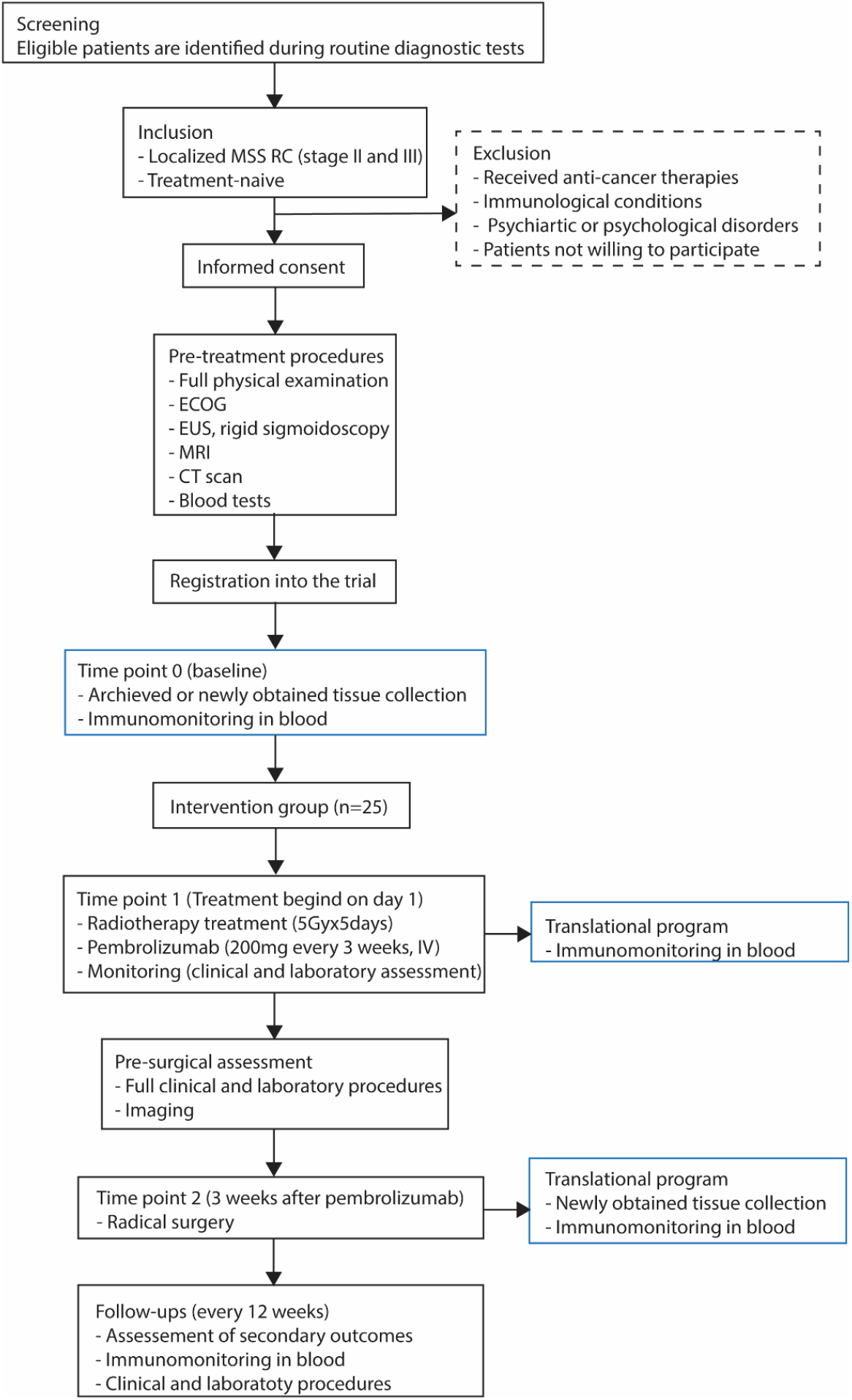
Flowchart of the study. RC: rectal cancer, MSS: microsatellite stable, ECOG: Eastern Co-operative Oncology Group, EUS: endoscopy ultrasound, MRI: magnetic resonance imaging, CT: computed tomography

### Study population

#### Inclusion criteria

Participants are eligible to be included in the study only if all of the following criteria apply:

1. Male/female participants must be at least 18 years old, have adequate health conditions and be willing to comply with the study protocol.
2. Participants must have been diagnosed with localized RC and not being treated for this disease before the start of the clinical trial.
3. A multi-disciplinary tumor board should recommend neo-adjuvant SCPRT and surgery as the optimal treatment option for these patients.

#### Exclusion criteria

Participants are excluded from the study if any of the following criteria apply:

1. Participants who have received anti-cancer therapy, live vaccine or have participated in other investigational trials in close proximity with the start of the clinical trial. If treated with RT, received major surgery or particpated in another study, the participant has to be fully recovered.
2. Participant suffering from active autoimmune diseases, infections, immunodeficiency, other malignancies or health conditions that could confound the results of the study and/or interfere with the subject’s participation for the full duration of the study.

### Hypothesis

We hypothesize that combining pembrolizumab with SCPRT can increase the TRG compared to the standard of care (RT alone) due to the increased immune infiltrate in the tumor (specifically CD8^+^ T cell and FOXP3^+^ T cell) in subject with localized MSS and MSI RC.

### Study interventions

The clinical trial will include the following therapeutic interventions:

#### Pembrolizumab

Four injections of pembrolizumab will be administrated intravenously (IV) at a fixed dose of 200mg, over a period of 9 weeks, starting on the first day of the SCPRT and repeated every three weeks. There is no maintenance treatment.

#### External beam radiotherapy

SCPRT will be administered using volumetric-modulated arc therapy (VMAT) to a prescription dose of 25 Gy in 5 fractions of 5 Gy to the planning target volume (PTV) over 5 days (Monday to Friday). RT starts on day 1 of treatment (always on a Monday).

#### Surgery

Surgical procedure will take place 12 weeks after the end of the RT, meaning three weeks after the last (fourth) injection of pembrolizumab.

An overview of the trial intervention is presented in **Table 1**.

**Table 1.**
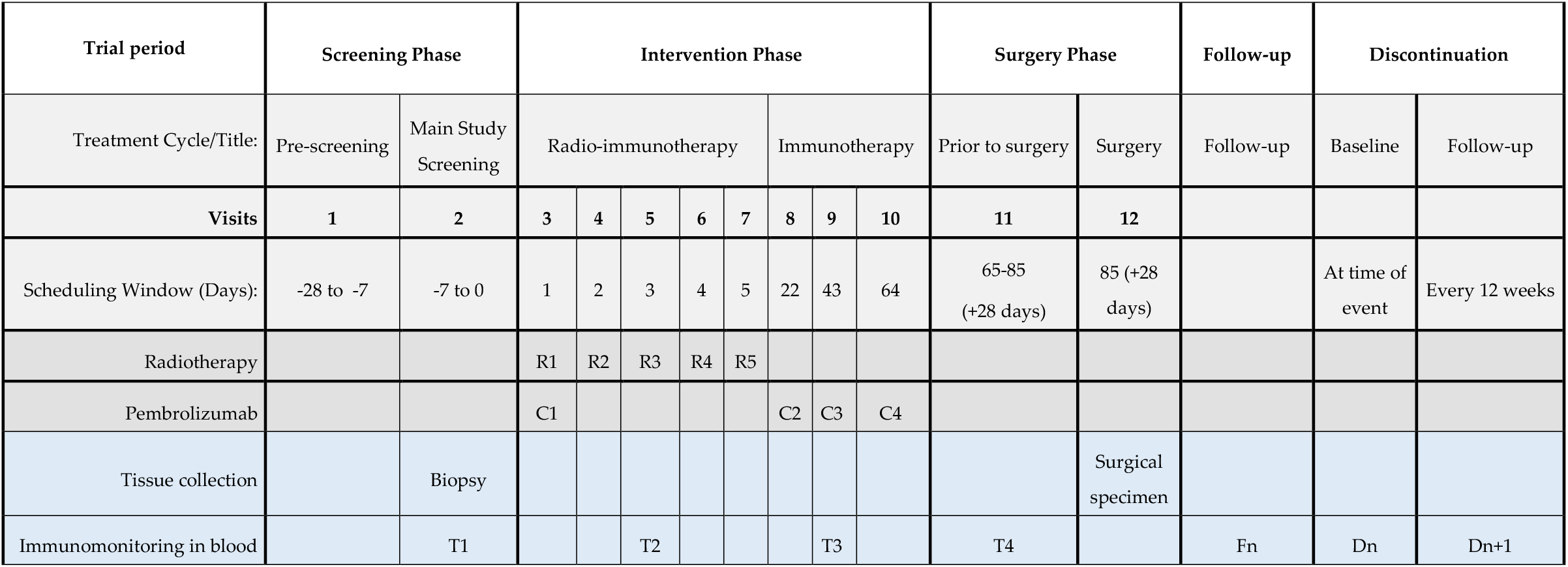
Trial interventions.

### Statistical analysis

The primary outcome of this study is TRG. The pathological complete response (pCR) rate, defined as the absence of residual invasive disease in the rectum and in the lymph nodes at the completion of the neoadjuvant treatment [24], reaches 12-16% in patients receiving treated with SCRT with delayed surgery or CRT, respectively [5]. Remarkably, induction CT before CRT and surgery increase pCR to 30%-35% [25]. Our hypothesis is that the abovementioned intervention results in a pCR rate that is superior to that of SCPRT. The null hypothesis assumes a 10% pCR rate, and the alternative a 30% pCR rate, which at the usual alpha (0.05) and beta (0.2) cutoffs, require a population of 25 patients, using an exact binomial sample size calculation. The null hypothesis will be rejected if at least 6 patients out of 25 meet the primary endpoint.

## Discussion

Currently, the standard of care for localized RC consists of neo-adjuvant treatments such as long-course CRT or SCPRT. Both treatments have been shown to increase local control and pCR compared to surgery alone. Contrary to dMRR/MSI-H tumors, novel cancer immunotherapies have shown little clinical impact on MSS CRC. Therefore, the Food and Drug Administration (FDA) granted approval for the use of such ICIs exclusively in the first- and second-line treatment of unresectable or metastatic dMMR/MSI-H CRCs. Remarkably, on top of the direct cytotoxic effects on tumor cells, RT is able to reprogram the TME to exert a potent anti-tumor immune response, thus providing a window of opportunity for immune-modulation. Recently, the first combined approach associating CRT with immunotherapy (Nivolumab) in the VOLTAGE trial [NCT02948348] was shown to be safe, and yielded promising results (30% pCR) in the treatment of MSS RC, but similar combinations (such as SCPRT with ICIs) have never been tested on RC.

This project investigates a novel combination treatment strategy for the treatment of localized RC. Specifically, we will study the clinical and biological impact of combining immunotherapy (pembrolizumab) with SCPRT in the neo-adjuvant treatment of localized MSS and MSI RC. This phase II non-randomized clinical trial is currently open and will recruit 25 patients that will be treated with SCPRT (5Gy x 5 consecutive days) and four injections of pembrolizumab followed by delayed surgery (NCT04109755). The study primary outcome is TRG, which is assessed by the Mandard regression grade score. Secondary outcomes include tolerability, safety, survival responses and immune and cellular dynamics in the tumor and in the peripheral blood. We postulate that RT will induce immunogenic cell death followed by inflammation, neoantigen release, resulting in elicitation of adaptive antitumor immune responses. These immune responses, mediated by tumor specific T cells, will be further amplified by the addition of the immune checkpoint inhibitor pembrolizumab. Upon combination treatment, we anticipate an increase of CD3^+^, CD8^+^ and FoxP3+ T cells in the TME, a dynamic change of the immune cell populations in the peripheral blood and an increased tumor regression as compared to historical controls due to the increased immune infiltrate within the tumor. Our clinical trial will generate important clinical data on the safety and efficacy of the combination and an invaluable insight into immunological changes linked to this combined modality.

## Data Availability

All data produced in the present study are available upon reasonable request to the authors.

## List of abbreviations

CRT: Chemoradiotherapy
CT: Chemotherapy
ctDNA: Circulating tumor DNA
CT: Computed tomography
CTLA-4: Cytotoxic T lymphocyte-associated protein 4
dMMR: Deficient mismatch repair mechanisms
DMFS: Distant metastasis-free survival
ECOG: Eastern co-operative oncology group
EUS: Endoscopy ultrasound
ICI: Immune checkpoint inhibitor
IV: Intravenously
LRRFS: Locoregional relapse-free survival
MRI: Magnetic resonance imaging
MSI: Microsatellite instability
MSS: Microsatellite stable
OS: Overall survival
PBMC: Peripheral blood mononuclear cell
PTV: Planning target volume
PD1: Programmed cell death 1
PD-L1: Programmed cell death ligand 1
PFS: Progression free survival
RT: Radiotherapy
RC: Rectal Cancer
Tregs: Regulatory T cells
SCPRT: Short-course preoperative radiotherapy
TME: Tumor microenvironment
TRG: Tumor regression grade
TIL: Tumor-infiltrating lymphocyte
VMAT: Volumetric-modulated arc therapy

## Declarations

### Ethics approval and consent to participate

This clinical study (registration number: NCT04109755) has received ethical approval from the Regional Research and Ethics Committee (CCER) and Swissethics (Protocol number: 2018-02346). A written informed consent is obtained from all participants upon participation.

### Consent for publication

Not applicable.

### Availability of data and materials

The datasets used and/or analyzed in the current study are available from the corresponding author on reasonable request.

### Authors’ contributions

Funding application: PYD and TK. Principal investigator: TK. Drafting of the Protocol Manuscript: CC. Intellectual Content: All authors. Study supervision: TK, CC. Revision and Final Approval of the Article: All authors.

## Acknowledgements

We would like to thank: The University Hospital of Geneva for supporting the study and all the patients for their contribution to the development of the intervention.

